# Apolipoprotein E-Genotyping and MRI Study for Alzheimer’s Disease Classification: PCR-RFLP and Restricted Enzymes AfIII for RS429358 and HaeII for RS7412

**DOI:** 10.1101/2024.01.04.24300735

**Authors:** NH Mohad Azmi, S Suppiah, NSN Ibrahim, B Ibrahim, VP Seriramulu, M Mohamad, T Karuppiah, NF Omar, N Ibrahim, RM Razali, NH Harrun, H Sallehuddin, N Syed Nasser, AD Piersson

## Abstract

The most common type of dementia in neurodegenerative diseases is Alzheimer’s disease (AD), a progressive neurological illness that causes memory loss. Neurophysiological tests, including the montreal cognitive assessment (MoCA), mini-mental state examination (MMSE), and clinical dementia rating (CDR) scores, are used to identify AD. Neuroimaging studies T1-weighted MRI scans assessed brain structural abnormalities. AD patients had grey matter volume (GMV) loss in brain structures when structural MRI data were analysed using voxel-based morphometry (VBM). Neuroimaging studies using resting state functional MRI (rs-fMRI)-blood oxygen level dependent (BOLD) sequence for brain imaging were processed using the seed-based analysis (SBA) method to analyse functional connectivity (FC) in the default mode network (DMN), sensorimotor network (SEN), executive control network (ECN), language network (LN), visuospatial network (VN), and salience network (SAN). Late-onset AD can be studied using the apolipoprotein E gene (ApoE). ApoE has four alleles with LOAD patients having either a homozygous or heterozygous genotype of these alleles. The genotypes, particularly ApoE ε4, are associated with a more significant risk for AD pathogenesis. The combination of genotyping and MRI neuroimaging is a promising avenue for research that starts with protocol optimisation. Objective: to differentiate changes in structural brain volumetric and rs-fMRI functional connectivity strength with the diagnosis of AD and HC by combining ApoE ε4 genetic variations.

**Materials and Methods:** Thirty participants with AD, n = 15, and healthy control (HC), n = 15, for the MRI study, and six participants (n = 6) with AD, n = 3, and HC, n = 3, for ApoE genotyping. In this study, we categorised the participants using neuropsychological tests, i.e., MoCA, MMSE, and CDR. We performed structural and functional MRI brain imaging to identify network areas affected by AD. Structural voxel-based morphometry (VBM) models and the CONN Toolbox, which analysed functional MRI using seed-based analysis (SBA), were performed. Genotyping was done by extracting the DNA from the participants’ blood samples. The isolated DNA underwent PCR-RFLP. Then, the restricted enzymes RE AFIII for rs429358 and HAEII for rs7412 were performed.

**Results:** There was decreased grey matter volume (GMV) and reduced functional connectivity among AD participants involving the frontal lobe and anterior cingulate gyrus in DMN, SEN, ECN, LN, VN, and SAN. We detected three participants with a homozygous ApoE ε4 negative genotype (non-carriers), which was consistent with the HC genotype. We also detected heterozygous genotype ApoE ε4 positive carriers, which indicated LOAD.

**Conclusion:** There is altered GMV in VBM, a decrease in brain activation, and an increase in spatial activation size in rs-fMRI neuronal FC in some areas of the brain with ApoE ε4 carriers in AD participants. Thus, the imaging features of the AD participants are well mapped to their ApoE ε4 carrier status. Thus, we propose our radiogenomics techniques as a useful biomarker for the characterisation of AD patients.

## Introduction

The most common type of dementia in neurodegenerative diseases is Alzheimer’s disease (AD), a progressive neurological illness that causes memory loss. Due to neuronal death, it starts with hippocampal shrinkage and extends to other parts of the brain (1, 2). One of the major ailments affecting the elderly is AD, whose incidence has dramatically increased recently (3). A previous study conducted among elderly Malaysians found that dementia was associated with elderly age, lack of formal education, gender, low self-rated health quality, and Malay or Bumiputera ethnicity (4, 5).

Neurophysiological tests, including the Montreal Cognitive Assessment (MoCA), Mini-Mental State Examination (MMSE), and Clinical Dementia Rating (CDR) Scores, are used to identify AD (6–8). The MoCA should not replace a more comprehensive neuropsychological examination (9–11). MMSE is helpful for screening dementia in older adults with basic literacy. However, it is likely to misclassify illiterate older adults, which has major consequences for AD detection in poor nations with low literacy rates (12). Dementia and cognitive change are assessed by the global dementia rating scale (CDR) (13). Thus, neuroimaging evidence supports Alzheimer’s diagnosis (14, 15).

Neuroimaging studies T1-weighted MRI scans assessed brain structural abnormalities (16–18). AD patients had grey matter volume (GMV) loss in brain structures when structural MRI data were analysed using voxel-based morphometry (VBM) (19–22). VBM, an MRI analysis method, can detect brain morphological abnormalities non-invasively (16, 23). VBM can compute neuronal cortices’ anatomical sections and quantify local brain tissue concentrations, mostly grey matter (24, 25) (26). AD risk was linked to grey matter atrophy (27). AD patients’ brain grey matter density (GMD) at each voxel can be compared (28, 29). In AD, researchers found that higher GMD was linked to lower GMV in the medial temporal lobe, the temporal gyri, the precuneus, the insular and cingulate cortex, and the caudate nucleus (29, 30).

Neuroimaging studies resting state functional MRI (rs-fMRI)-blood oxygen level dependent (BOLD) sequence for brain imaging was processed using the seed-based analysis (SBA) method to analyse functional connectivity (FC) (31). SBA can evaluate functional changes between participants, analyse a region of interest (ROI), and show the FC among different brain networks such as default mode network (DMN) (32, 33), sensorimotor network (SEN) (34), executive control network (ECN) (35), language network (LN) (36), visuospatial network (VN) (37), and salience network (SAN) (38). Historically, DMN, SEN, ECN, LN, VN, and SAN are the networks that have a characteristic pattern of FC on rs-fMRI. FC has been implicated in altered brain morphometry in various neurological and psychological disorders, including AD (38, 39). An AD brain investigation found reduced FC, notably in DMN nodes (40), SEN nodes (41), ECN nodes (42), LN nodes (43), and VN nodes (43) compared to HC patients. Despite many studies conducted in Caucasian and North Asian populations, there is a lack of data regarding rs-fMRI in Malaysia to elucidate the changes that occur in AD as well as identify imaging biomarkers in our population.

Genotyping ApoE involves comparing an individual’s DNA sequence to another or a reference sequence to find genetic differences. For our participants, we study ApoE ε2, ε3, and ε4 alleles. The ApoE ε4 allele is a risk factor for LOAD-type Alzheimer’s disease (35, 44–46). Mammals use ApoE to metabolise lipids, while humans encode it for diverse roles (47). Researchers can study population genetic diversity and evolution by analyzing haplotypes. LOAD development has been linked to a positive homozygous or heterozygous ε4 subtype (48). ApoE ε4 inheritance patterns include negative (no allele), heterozygous (one allele), or homozygous (two alleles). The homozygous ε4 allele is associated with the highest risk of LOAD pathology (46, 47). It promotes amyloid deposition and causes neuronal cell inflammation. Over-60s are at higher risk for LOAD due to gene mutations in 95% of cases (43). Early-onset AD affects 2% to 3% of people under 60 (49). Genotyping can be done using PCR-RFLP and RE (37). ApoE data in Malaysian AD patients is scarce.

We hypothesize that there will be an association between altered GMV in T1 VBM, a decrease in brain activation, and an increase in spatial activation size in rs-fMRI neuronal FC in some areas of the brain and the ApoE ε4 of AD participants. In this study, we used ApoE ε4 genetic variations to look at changes in structural brain volumetric and rs-fMRI functional connectivity strength in people who had been diagnosed with AD and HC.

## Materials And Methods

Thirty participants (n = 30) with AD, n = 15, and healthy control (HC), n = 15, for the MRI study, and six participants (n = 6) with AD, n = 3, and HC, n = 3, for ApoE genotyping. In this study, we categorised the participants using neuropsychological tests, i.e., MoCA, MMSE, and CDR.

### A. Study design and participant recruitment

This prospective cross-sectional study was approved by the institutional ethical committee (JKEUPM-2019-328) and MREC (NMRR-19-2719-49105). This study collected data from March 2021 until June 2022. We surveyed AD patients from Hospital Kuala Lumpur (HKL) memory clinic, Klinik Kesihatan Pandamaran Klang (KKP), and Hospital Pengajar UPM (HPUPM) to recruit eligible Klang Valley AD volunteers for this study. Cognitively healthy, age-matched controls were selected. This study included only eligible participants. Prior to recruitment, potential participants gave informed consent to participate in the study, following the Declaration of Helsinki. All data were anonymised and participants were paid. We compensated the participants and anonymized all data.

### B. Inclusion and exclusion criteria

Participants must have a clinically verified AD diagnosis and be 55–90 years old. Additionally, participants must be Malaysians. Researchers divided participants into AD and healthy control HC groups based on their DSM-5, MoCA, MMSE, and CDR scores. HC group members had to be cognitively strong and free of neurological diseases like cancer or stroke. The participants did not have metal implants, did not have claustrophobia, and cooperated throughout resting-state functional magnetic resonance imaging (rs-fMRI). The study excluded non-Malaysians and those with neurological illnesses other than Alzheimer’s. Claustrophobia, irremovable metallic implants, electronic implants including pacemakers, cochlear or ear implants, and metallic tattoos were relative and absolute contraindications for magnetic resonance imaging (MRI).

### C. Alzheimer’s disease and neurophysiological assessment

The patients underwent the administration of various questionnaires, including the Montreal Cognitive Assessment (MoCA), Mini-Mental State Examination (MMSE), and Clinical Dementia Rating (CDR) scores.

#### C.I. Montreal Cognitive Assessment (MoCA)

We assessed short-term memory using an 8-item self-reported MoCA questionnaire. Executive functions, visuospatial ability, attention, concentration, working memory, language, and time/place orientation were assessed (MoCA Cognitive Assessment). For visuospatial ability, clock-drawing scores 3 points and three-dimensional cube copies 1 point. Short-term memory recall scores 5 points for two learning trials of five nouns and delayed recall after five minutes. The examiner administered a verbal language test. One point was given for sustained attention, focus, and working memory; three points for serial number subtraction; and one point for digit span forward and backwards. The assessment of language includes a three-item confrontation naming test with low-familiar animals (lion, camel, rhinoceros), repetition of two syntactically complex sentences for 2 points, and fluency. Finally, participants score 6 points for orientation to time and place by providing the exam date and city. MoCA scores are 0–30. A normal MoCA questionnaire score is 26 or higher. Research showed that people without cognitive impairment scored 27.4, MCI scored 22.1, and Alzheimer’s disease scored 16.2 (50–52).

#### C.II. Screening tool: The Mini-Mental State Examination (MMSE)

Five-item self-reported MMSE or Folstein tests are 30-point questionnaires. The questionnaire was screened for dementia. Ten points for orientation (time and place), three for registration. 5 points for attention and computation, 3 for recall, and 9 for language (repetition, complex commands). Participants scoring <26 for Alzheimer’s disease and ≥26 were deemed normal after assessing the MMSE questionnaire (53, 54).

#### C.III. Clinical Dementia Rating (CDR)

Researchers worldwide use CDR to assess dementia severity. Morris (1993) developed a numerical dementia severity scale. A qualified medical or psychological staff will use Charles Hughes’ 1982 structured interview protocol to assess a patient’s cognitive and functional performance in six cognitive areas: orientation, memory, judgement and problem-solving, home and hobbies, community affairs, and personal care. Each of these results is added for a 0–3 composite score. This score helps characterise and track a patient’s dementia or impairment. 0 = normal, 0.5 = very mild, 1 = mild, 2 = moderate, and 3 = severe (55, 56).

### D. Pre-processing Structural Brain Volumetric using Voxel-Based Morphometry (VBM) Analysis

We conducted the magnetic resonance imaging (MRI) procedure using a Siemens 3.0T scanner, specifically the PRISMA model manufactured by Siemens in Erlangen, Germany. A structural MRI was conducted using a 12-channel head coil. High-resolution T1-weighted magnetization We obtained high-resolution T1-weighted Magnetization Prepared Rapid Gradient Echo (MPRAGE) magnetic resonance imaging (MRI) data. The parameters of the sequence were as follows: The parameters used for the imaging sequence were as follows: repetition time (TR) of 2300 milliseconds, echo time (TE) of 2.27 milliseconds, and inversion time (TI) of 1100 milliseconds, for a total of 160 slices acquired in an ascending sagittal orientation. The imaging sequence used a field of view (FOV) of 256 x 256 mm2, with a matrix size of 256 x 256. The slice thickness was set to 1mm, ensuring high-resolution imaging. The imaging method used a T1-weighted magnetization-prepared rapid gradient echo pulse sequence with 160 slices, a repetition time (TR) of 2300 milliseconds, an echo time (TE) of 2.27 milliseconds, and an inversion time (TI) of 1100 milliseconds.

We used the VBM toolbox in the Statistical Parametric Mapping software (SPM 12, http://www.fil.ion.ucl.ac.uk/spm/software/spm12) to pre-process structural images (22, 57, 58)

Initially, all images were checked for artefacts and structural abnormalities. Secondly, the temporal processing encompassed slice timing, while thirdly, the spatial processing involved realignment and estimation, setting the origin, co-registration, normalising, and smoothing. The researchers employed the group-specific AD and HC templates in order to minimise heterogeneity across the subjects. They subsequently used the "DARTEL Normalize to Montreal Neurological Institute MNI Space" program to standardize the images with the Asian brain map template. We determined the volume of a particular region of interest (ROI) by spatially aligning T1-weighted images to the MNI template, relying on pre-existing knowledge. The measurement of volume changes was conducted by retaining segmented pictures of the GMV, which were obtained through spatial registration and modulation of the images to reflect tissue volumes. Subsequently, a Gaussian filter with a full width at half maximum (FWHM) of 8mm was applied to the normalised brain images in order to achieve smoothing. We employed the family-wise error (FWE) to account for multiple comparisons at a significance level of p<0.05. In order to identify regions exhibiting weak signals, we reduced the threshold utilized in the SPM analysis, which was considered uncorrected for family-wise error (FWE), to a significance level of p<0.001.

### E. Pre-processing Resting-state MRI (Rs-MRI) using seed-based analysis

We performed functional imaging using an echo-planar imaging (EPI) sequence. The phase encoding direction employed in the study was from the anterior to posterior directions. The imaging parameters used were as follows: repetition time (TR) of 3000 ms, echo time (TE) of 30 ms, slice thickness of 3mm, field of view (FOV) measuring 220 × 220 mm2, voxel size of 2.3 x 2.3 x 3mm, and a total of 38 slices were acquired. The participants were instructed to recline in a supine position with their eyes shut while being advised to refrain from entering a state of sleep.

We conducted the study of resting-state functional connectivity (rs-FC) using the CONN toolbox v20.b (http://www.nitrc.org/projects/conn) with a seed-based approach. We subsequently conducted seed-based analysis (SBA) utilizing regions of interest (ROIs) established based on pre-existing knowledge, following a comprehensive investigation of the whole brain. The SPM12 software was used to pre-process the functional images. This included a lot of steps, such as fixing the slice timing, realigning the images, registering them with the T1-weighted anatomical image, normalising the images to the MNI space, and smoothing (54). We established the significance threshold for this study at p<0.05, using the cluster-size p-FDR corrected p<0.05 approach for family-wise error controls.

### F. ApoE Genotyping

Genotyping was done by extracting the DNA from a patient’s blood sample. We checked the quality of the isolated DNA using a nanodrop spectrophotometer. Participants whose DNA quality was good enough went through polymerase chain reaction-based restriction fragment length polymorphism (PCR-RFLP). A PCR-RFLP assay detected the alleles of rs429358 and rs7412, respectively. The amplication reaction system contained 100 g of DNA template (volume: 17µl) added to 25µl of MyFi Mix and 1µl primer. Each allele got 50µl of nuclease-free water added to it. For rs429358, the forward primer was AGGGCGCTGATGGACGAGAC and the reverse primer was GCCCCGGCCTGGTACACT. For rs7412, the forward primer was GGCGCGGACATGGAGGAC, and the reverse primer was GCCCCGGCCTGGTACACT. We set the PCR cycling conditions as follows: Initial denaturation (1) at 95°C for 3 minutes [1 cycle]. Denaturation (2): 95°C for 30 seconds [35 cycles]. Annealing (3) at 95°C for 30 sec [35 cycles]. Final extension: (4) 72°C for 5 minutes [1 cycle]. Hold: 4 °C. Digest 1µg of the amplified product with 1µl of restriction endonuclease (AflIII for rs429358 and HaeII for rs7412), 5µl of buffer, and 50µl of nuclease-free water. Mix the reaction by pipetting up and down and microfuging briefly. Incubate at 37°C for 60 minutes (rs429358) or 5–15 minutes (rs7412). Heat inactivates at 80°C for 20 minutes. Finally, we diluted the digested product tenfold and analysed it by capillary electrophoresis to detect the alleles of rs429358 and rs7412. The haplotypes of rs429358 and rs7412 determined the ApoE genotype. The ApoE ε2/ε2 allele was recognised by rs429358-TT and rs7412-TT (no risk of AD), the ApoE ε2/ε3 allele was identified by rs429358-TT and rs7412-TC (early onset of AD), the ApoE ε2/ε4 allele was identified by rs429358-TC and rs7412-TC (late onset of AD), the ApoE ε3/ε3 allele was identified by rs429358-TT and rs7412-CC (reduce risk of AD), the ApoE ε3/ε4 allele was identified by rs429358-TC and rs7412-CC (reduce risk of AD), and the ApoE ε4/ ε4 allele was defined by rs429358-CC and rs7412-CC (risk of AD). The ApoE ε4 inheritance pattern includes ApoE ε4 negative individuals as ε4 non-carriers, and ε4 carriers with a heterozygous genotype (TC) or a homozygous genotype (CC and TT) for the ApoE ε4 allele.

### G. Statistical Analysis

SPM 12 and Statistical Package for the Social Sciences (SPSS software Version 25.0, SPSS Inc., Chicago, IL, USA) were used for statistical analysis. We compared the demographic data and neuropsychological test scores between AD and HC using SPSS’s chi-square test and independent t-test. For T1 structural MRI analysis of VBM, the two-sample t-test was used to compare differences in brain GMV. We set the significance threshold at a p-value of less than 0.05. Regression analysis in ROI-based analysis of default mode network (DMN), sensorimotor network (SEN), executive control network (ECN), language network (LN), visuospatial network (VN), and salience network (SAN) data to identify regions of brain activation, specifically functional connectivity and spatial activation size, in both participants using SBA of rs-fMRI data. We set the significance level at a voxel threshold p-value of less than 0.05 and a cluster threshold p-value of less than 0.05.

## Results

### A. Demographic characteristics

We recruited thirty individuals. We dichotomised the participants into two categories: fifteen individuals from each category, i.e., fifteen AD and fifteen HC. The average age of the AD and HC populations was 70.6± 8.55 (59–83) and 69.27±6.81 (60–82) years, respectively. The gender distribution in the AD and HC groups was 5 males (33.3%) and 10 females (66.7%) in both groups. The educational backgrounds of participants varied, for AD and HC, with less than six years 3 (20.00%) and more than six years 12 (80.00%) and, less than six years 1 (6.67%) and more than six years 14 (93.33%) respectively. The marital status of participants varied, with single 1 (6.67%) and married 14 (93.33%), for AD and single 2 (13.33%) and married 13 (26.67%) for HC. These demographic data provide an overview of the characteristics of our study population. It is important to note that the groups were matched as closely as possible to ensure that any observed differences between the AD group and the HC group can be attributed to the condition under investigation rather than demographic factors (see table 1).

**Table 1:** Comparison of the sociodemographic and neuropsychological profiles of AD and HC.

| Variable<br>*n*=30 | | AD (*n*=15)<br>Freq. (%) | HC (*n*=15)<br>Freq. (%) | $X^2$ statistic<sup>a</sup> (df)<br>P Value<sup>a</sup> |
| --- | --- | --- | --- | --- |
| Demographic Data | **Gender** |  |  | 0.001 (1) 1.0 |
|  | Male | 5 (33.3%) | 5 (33.3%) |  |
|  | Female | 10 (66.7%) | 10 (66.7%) |  |
|  | **Education Level** |  |  | 1.15 (1) 0.28 |
|  | <6 years | 3 (20.00%) | 1 (6.67%) |  |
|  | >6 years | 12 (80.00%) | 14 (93.33%) |  |
|  | **Marital Status** |  |  | 0.37 (1) 0.54 |
|  | Single | 1 (6.67%) | 2 (13.33%) |  |
|  | Married | 14 (93.33%) | 13 (26.67%) |  |
|  |  | **Min-max**<br>**(mean ± SD)** | **Min-max**<br>**(mean ± SD)** | *t* statistic<sup>b</sup> (df)<br>P Value<sup>b</sup> |
|  | **n=30**<br>**Age Neuroimaging** | **n=15**<br>**59-83**<br>**(70.6 ± 8.55)** | **n=15**<br>**60-82**<br>**(69.27 ± 6.81)** | -0.47 (28) 0.64 |
|  | **n=6**<br>**Age Genotyping** | **n=3**<br>**61-83**<br>**(75.67 ± 12.7)** | **n=3**<br>**60-79**<br>**(68 ± 9.85)** | -0.83 (3.77) 0.46 |
| Neuropsychologic<br>al test scores. | **MoCA** | 0-22<br>(14.13 ± 6.82) | 23-30<br>(28.13 ± 2.20) | 7.56 (16.88)<br>0.001 |
| | **MMSE** | 0-29<br>(14.40 ± 8.33) | 24-30<br>(28.00 ± 2.17) | -13.26 (14.00)<br>0.001 |
| | **CDR** | 1-3<br>(2.13 ± 0.92) | 0<br>(0.001 ± 0.001) | 6.12 (15.89)<br>0.001 |

**Note:** AD: Alzheimer's disease group; HC: healthy control group; MoCA: Montreal Cognitive Assessment; MMSE: Mini-Mental State Examination; CDR: Clinical Dementia Rating Score
<sup>a</sup> Chi-square test for independence (*n*= Frequency *df*= Degree of Freedom) with participant categories AD and HC.
<sup>b</sup> independent *t* test for independence (*n*= Frequency *df*= Degree of Freedom) with participant categories AD and HC.

We recruited six individuals. We dichotomised the participants into two categories: three individuals from each category, i.e., three AD APOE ε4 carriers and three HC APOE ε4 non-carriers. The average age of the AD and HC populations was 75.67±12.7 (61–83) and 68±9.85 (60–79) years, respectively (see table 1).

### B. Neuropsychological assessment test

The MoCA, MMSE, and CDR scores were assessed, and the results are shown in Table 1.

### C. Voxel-Based Morphometry (VBM) analysis

In the whole brain analysis of participants with contrast AD more than HC to detect the brain density, there were signals at (1) left anterior orbital gyrus (25%), left lateral orbital gyrus (20%), left middle frontal gyrus (15%), left frontal lobe (1%), left superior frontal gyrus (0.2%) in coordinates −32 +54 −9, (2) putamen (47%) in coordinates −26 +12 −3, (3) right caudate (14%), right putamen (12%), right medial orbital gyrus (4%), right anterior cingulate gyrus (1%), right gyrus rectus (1%), right medial frontal cortex (0.6%) in coordinates 12 +16 −9, (4) left medial orbital gyrus (41%), left posterior orbital gyrus (7%), left gyrus rectus (6%), Left medial frontal cortex (0.2%) in coordinates −16, 22, and 20, and (5) the left superior frontal gyrus medial segment (20%) and the left superior frontal gyrus (17%) in coordinates −12 + 34 + 39 were found to have high grey matter density or voxel density for AD compared to the HC group (uncorrected p-value <0.001). (47%) in coordinates −26 +12 −3, found to be high grey matter density or voxel density for AD compared to the HC group (uncorrected family-wise error (FWE p-value <0.001), indicative of increased density but reduced GMV of these regions in the AD participants. Hence, a significant reduction in GMV was noted in the AD compared to HC participants using T1 MRI structural data (Table 2 and Figure 1).

**Figure 1.**
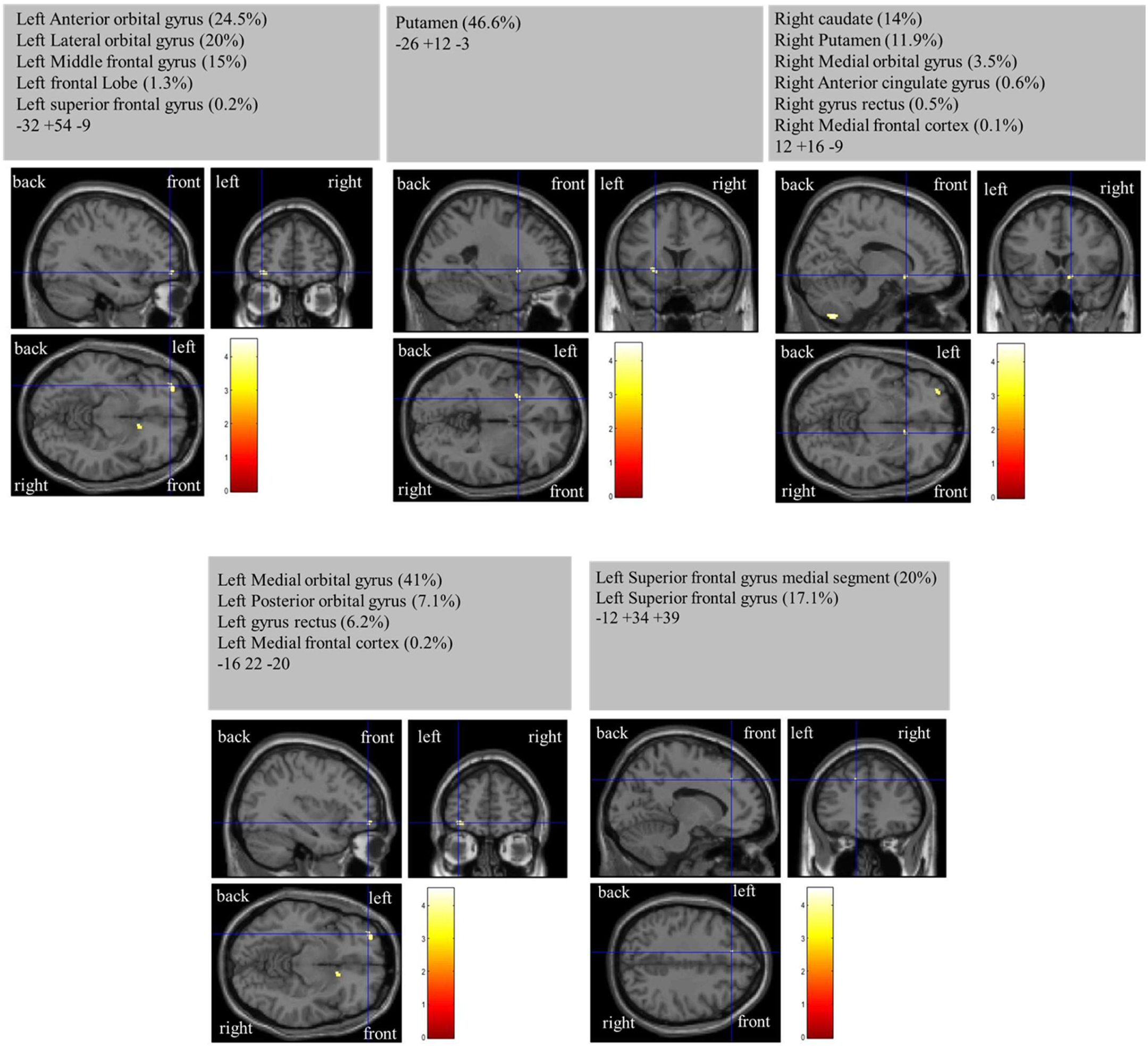
VBM results had different density and reduced grey matter volume in AD participants more than HC participants using T1 MRI structural data, uncorrected p-value 0.001 images (p<0.05, FWE corrected).

**Table 2:** Tabulated values of regional differences in voxel density for AD > HC group and HC > AD images (uncorrected family-wise error (FWE p-value <0.001))

| AD>HC |  | Side | Cluster<br>Peak<br>mm mm<br>mm | Voxel | Peak T |  | P Level<br>(FWE) |
| --- | --- | --- | --- | --- | --- | --- | --- |
| Anterior orbital gyrus | aOrG Lt | Left | -32 +54 -9 | 56 | 4.04 | 24.6% | <0.001 |
| Lateral orbital gyrus | LorG Lt | Left | -32 +54 -9 | 56 | 4.04 | 20.0% | <0.001 |
| Middle frontal gyrus | MFG Lt | Left | -32 +54 -9 | 56 | 4.04 | 15.0% | <0.001 |
| Frontal lobe | FL Lt | Left | -32 +54 -9 | 56 | 4.04 | 1.3% | <0.001 |
| Superior frontal gyrus | SFG Lt | Left | -32 +54 -9 | 56 | 4.04 | 0.2% | <0.001 |
| Putamen | Putamen Lt | Left | -26 +12 -3 | 39 | 4.5 | 46.6% | <0.001 |
| Caudate | Caudate Rt | Right | 12 +16 -9 | 26 | 3.73 | 14.0% | <0.001 |
| Putamen | Putamen<br>Rt | Right | 12 +16 -9 | 26 | 3.73 | 11.9% | <0.001 |
| Medial orbital gyrus | MOrG Rt | Right | 12 +16 -9 | 26 | 3.73 | 3.5% | <0.001 |
| Anterior cingulate gyrus | ACgG Rt | Right | 12 +16 -9 | 26 | 3.73 | 0.6% | <0.001 |
| gyrus rectus | gyrus<br>rectus Rt | Right | 12 +16 -9 | 26 | 3.73 | 0.5% | <0.001 |
| Medial frontal cortex | MFC Rt | Right | 12 +16 -9 | 26 | 3.73 | 0.1% | <0.001 |
| Medial orbital gyrus | MOrG Lt | Left | -16 22 -20 | 3 | 3.45 | 41.0% | <0.001 |
| Posterior orbital gyrus | pOrG Lt | Left | -16 22 -20 | 3 | 3.45 | 7.1% | <0.001 |
| gyrus rectus | gyrus<br>rectus Lt | Left | -16 22 -20 | 3 | 3.45 | 6.2% | <0.001 |
| Medial frontal cortex | MFC Lt | Left | -16 22 -20 | 3 | 3.45 | 0.2% | <0.001 |
| Superior frontal gyrus<br>medial segment | mSFG Lt | Left | -12 +34 +39 | 6 | 3.19 | 20.0% | <0.001 |
| Superior frontal gyrus | SFG Lt | Left | -12 +34 +39 | 6 | 3.19 | 17.1% | <0.001 |
| AD<HC |  | Side | Cluster<br>Peak | Voxel | Peak T |  | P Level<br>(FWE) |
| - |  | - | - | - | - |  | - |

**Note:** AD: Alzheimer's disease group; HC: healthy control group

### D. Seed-based analysis (SBA)

In Figure 2, brain regions that showed significant FC differences within the seed between AD and HC participants are demonstrated. Using SBA, neural FC showed reduced activation values in the DMN, SEN, ECN, LN, VN, and SAN among AD participants.

**Figure 2.**
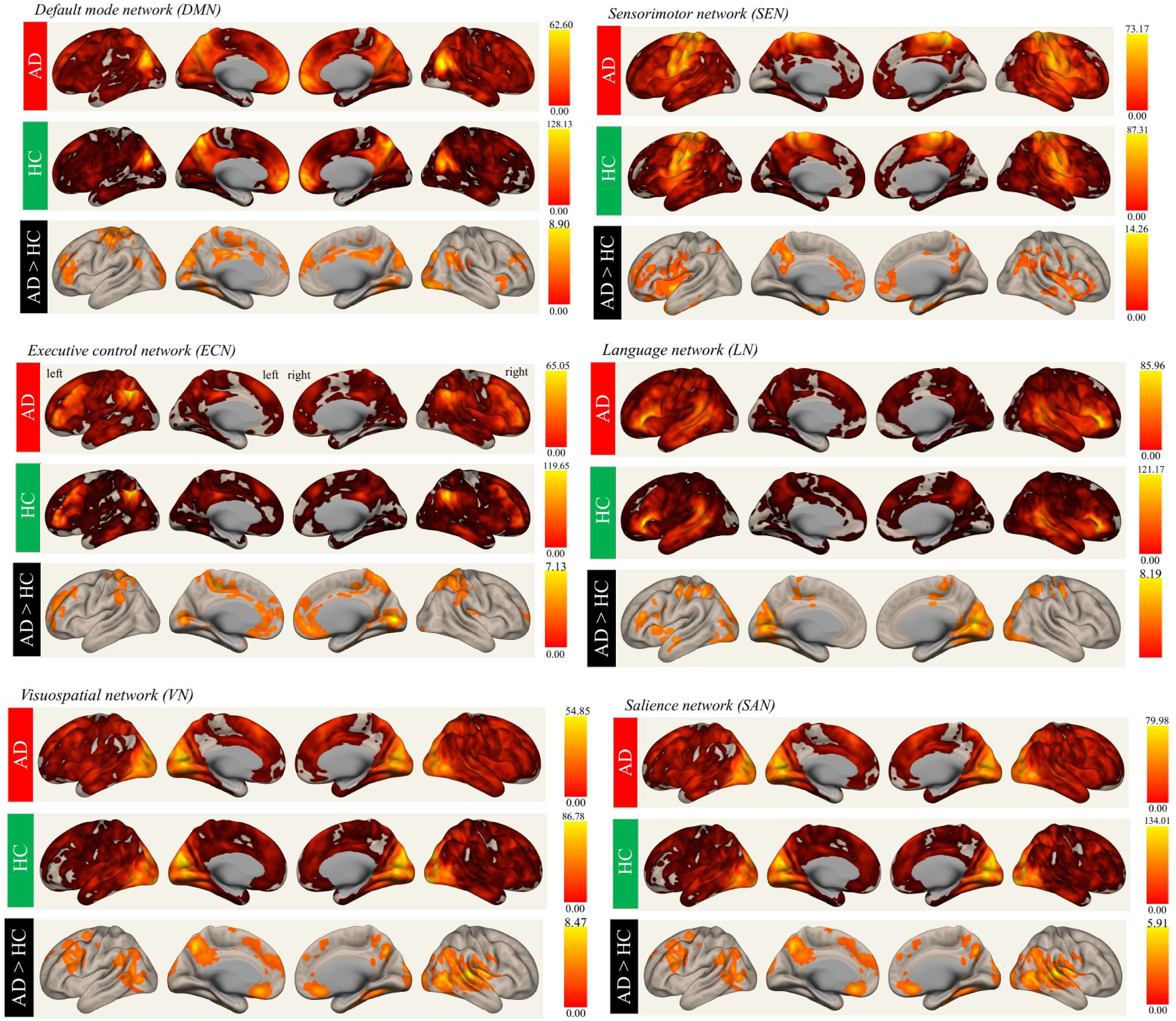
FC analysis for the LOAD and HC groups shows selectively rendered images for the DMN, SEN ECN, LN, VN, and SAN. Activation maps are graphical representations of activation regions in the brain. Where hot colours represent greater mean regional activation between the specific region and cold colours represent lower activation group differences. Activation values are based on T values. (FC: functional connectivity, DMN: default mode network, SEN: sensorimotor network, ECN: executive control network, LN: language network, VN: visuospatial network, and SAN: sailence network)

The findings of the study indicate a noticeable augmentation in the spatial activation size within the left postcentral gyrus (PostCG Lt), a component of the DMN, with a cluster size of 1389 voxels. Simultaneously, there is an observed augmentation in the spatial activation size inside the left frontal lobe (FL Lt) in the brain networks referred to as the DMN, SEN, and ECN, with respective cluster sizes of 1058, 990, and 1097 voxels. There is an observed augmentation in the spatial activation size inside the precuneous (PCC) region of the brain networks referred to as SEN, VN, and SAN, with respective cluster sizes of 885, 1447, and 988 voxels. There was an observed augmentation in the spatial activation size located in the right occipital region, specifically in the area known as Pole OP Rt. This increase was observed inside the brain networks referred to as LN and VN, with corresponding cluster sizes of 1081 and 971 voxels, as indicated in Table 3.

**Table 3:**
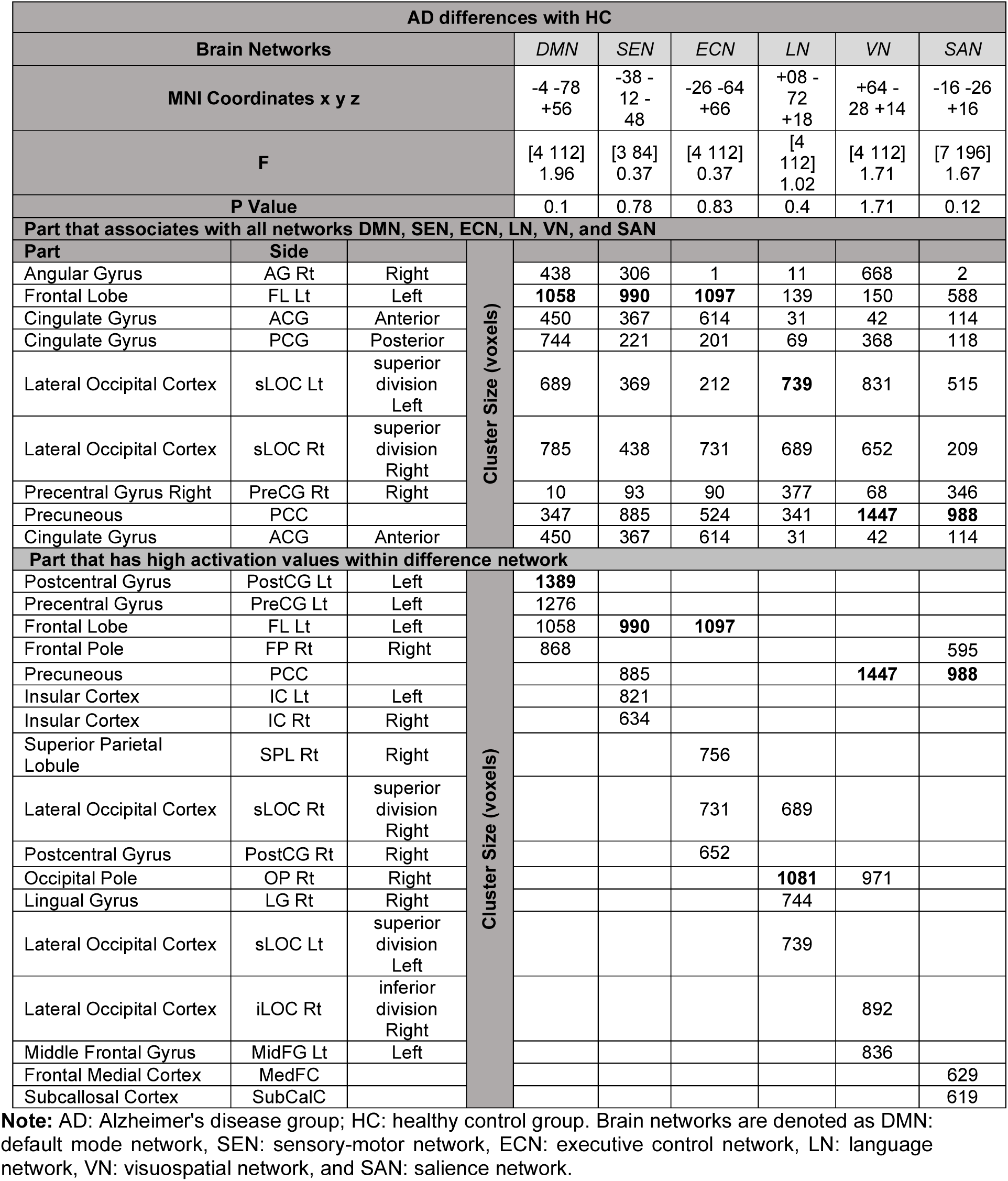
Tabulated regional differences in seed-based rs-fMRI functional connectivity found in all brain networks: DMN, SEN, ECN, LN, VN, and SAN, and high activation values in different brain networks. Rs-FC analysis indicated a significance level of p<0.05, with a cluster-size p-FDR corrected to p<0.05.

A reduction in activation values was noted in individuals diagnosed with AD in comparison to HC across various brain regions. These regions include the right angular gyrus (AG Rt), left frontal lobe (FL Lt), anterior cingulate gyrus (ACG), posterior cingulate gyrus (PCG), lateral occipital cortex, superior division left (sLOC Lt), lateral occipital cortex, superior division right (sLOC Rt), precentral gyrus right (PreCG Rt), and precuneous (PCC) within all brain networks. The decrease in activation was detected in all brain networks, including the DMN, SEN, ECN, LN, VN and SAN (refer to figure 2).

### D. ApoE Genotyping

In our investigation, the ApoE allele variants observed were ε2/ε3, rs429358 (TT), and rs7412 (CC) among individuals who do not carry the ε4 allele of ApoE. For individuals classified as HC with a non-carrier status for the ε4 allele, the inheritance pattern revealed that all three patients were homozygous for the aforementioned variants. The ApoE allele variants include ε2/ε4, rs429358 (TC), and rs7412 (TC) for individuals with AD who carry the ε4 allele. Inheritance patterns for AD ε4 carriers indicate that they are heterozygous for all three participants (Figure 3).

**Figure 3.**
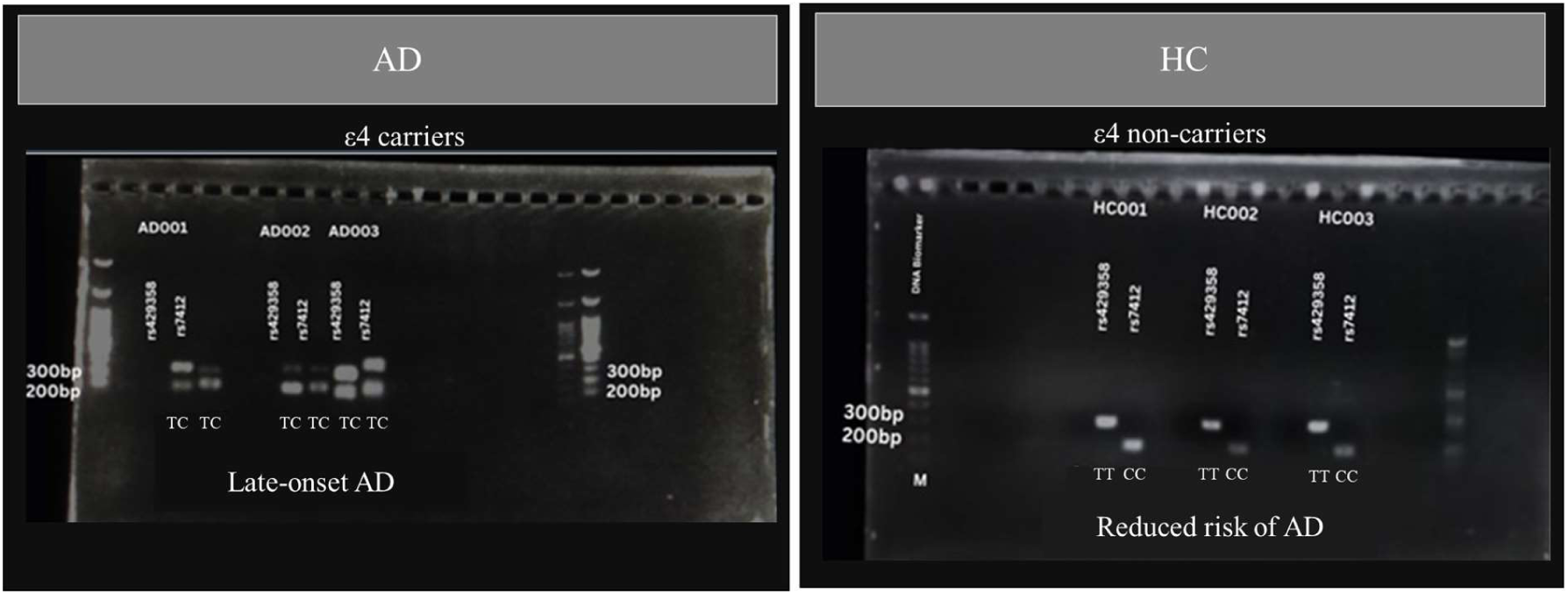
Agarose gel electrophoresis (2%) stained with ethidium bromide shows the genotype of the PCR products after digestion by restriction endonucleases. HC001, HC002, and HC003: TT, CC homozygous genotype, AD001, AD002 and AD003: TC, TC heterozygous genotype M: DNA Biomarker Note: AD: Alzheimer’s disease group, HC: healthy control group.

## Discussion

In our findings the MoCA (59), MMSE (12), and CDR scores (60) between AD and HC are significant differences (p<0.001). The scores identified the AD and HC categories.

In our neuroimaging studies T1-weighted MRI scans, we found altered GMV in VBM associated with specific regions of the AD brain, part of the frontal lobe namely the left anterior orbital gyrus (aOrG Lt), left posterior orbital gyrus (pOrG Lt), left lateral orbital gyrus (LorG Lt), left medial orbital gyrus (MOrG Lt), right medial orbital gyrus (MOrG Rt), left gyrus rectus, right gyrus rectus, left superior frontal gyrus (SFG Lt), left middle frontal gyrus (MFG Lt), right medial frontal cortex (MFC Rt), left medial frontal cortex (MFC Lt), left superior frontal gyrus medial segment (mSFG Lt), left superior frontal gyrus (SFG Lt), left frontal lobe (FL Lt), the frontal lobe area affected the decision-making, problem-solving, planning, and controlling emotions. It also plays a role in personality expression, social behaviour, and motor movements (61); the right anterior cingulate gyrus (ACgG Rt) It is involved in certain higher-level functions, such as attention allocation, reward anticipation, decision-making, ethics and morality, impulse control, and emotion (62); The left putamen (Putamen Lt), right putamen, and right caudate (Caudate Rt) (63) help regulate and fine-tune motor movements, and they also help in storing and retrieving information about motor patterns and habits, as evidenced by decreased GMV among the AD participants, thus resulting in loss of attention and memory impairment, respectively, as have been postulated by *a priori* knowledge shared by Japee et al. and Wang et al. (22, 64).

Our study revealed that individuals with Alzheimer’s disease (AD) exhibited atrophy in the grey matter volume (GMV) and a reduction in the size of spatial activation in resting-state functional connectivity (rs-FC) across all brain networks in the left frontal lobe (FL Lt) (65) and right anterior cingulate gyrus (ACG Rt) (66). We observed these findings in both voxel-based morphometry (VBM) and surface-based analysis (SBA). Utilising our restricted enzyme technique, we also found a strong link between the Apolipoprotein E ε4 (ApoE ε4) allele being present in AD patients (37, 44). These findings were in contrast to the healthy control (HC) group, where individuals’ non-carriers of the ApoE ε4 allele were identified (32, 36, 38, 67).

Furthermore, the resting-state FC was noted to be reduced in activation for AD in all brain networks, i.e., DMN, SEN, ECN, LN, VN, and SAN, which was counterintuitive to our hypothesis as we expected reduced functioning in these regions. Nevertheless, our findings corroborate the postulations by Mevel et al. that stated increased FC activity in the frontal DMN may be a reflection of the compensatory mechanism to overcome decreased FC at the posterior DMN that has mainly affected the posterior cingulate cortex (68).

## Conclusion

In conclusion, the severity of the impaired FC of the resting-state networks is seen in the regions that have been robustly analysed for the presence of GMV atrophy. Additionally, we can accurately map the imaging features of the AD participants to their ApoE ε4 carrier status. Thus, we propose our radiogenomics techniques as a useful biomarker for the characterisation of LOAD patients.

## Data Availability

All data produced in the present study are available upon reasonable request to the authors

## Acknowledgements

The Malaysian Ministry of Higher Education awarded grant number 5540244 from the Fundamental Research Grant Scheme (FRGS 06-02-14-1497FR/5524581) to fund this research. We also express our gratitude to the Ministry of Health Malaysia and the Malaysian Society of Radiographers for their unwavering support of this research. We are also grateful to the personnel at Pusat Pengimejan Diagnostik Nuklear, UPM, who contributed directly or indirectly to the data collection.

## References

1. Piersson AD, Ibrahim B, Suppiah S, Mohamad M, Hassan HA, Omar NF, et al. Multiparametric MRI for the improved diagnostic accuracy of Alzheimer’s disease and mild cognitive impairment: Research protocol of a case-control study design. PLoS One. 2021;16(9):e0252883.

2. Ribeiro LG, Busatto GF. Voxel-based morphometry in Alzheimers disease and mild cognitive impairment: Systematic review of studies addressing the frontal lobe. Dement Neuropsychol. 2016;10(2):104–12.

3. Azmi M, Saripan M, Nordin A, Saad FA, Aziz SA, Adnan WW, et al. 18F-FDG PET brain images as features for Alzheimer classification. Radiation Physics and Chemistry. 2017;137:135–43.

4. Ibrahim B, Suppiah S, Ibrahim N, Mohamad M, Hassan HA, Nasser NS, et al. Diagnostic power of resting-state fMRI for detection of network connectivity in Alzheimer’s disease and mild cognitive impairment: A systematic review. Hum Brain Mapp. 2021;42(9):2941–68.

5. Szabo-Reed AN, Vidoni E, Binder EF, Burns J, Cullum CM, Gahan WP, et al. Rationale and methods for a multicenter clinical trial assessing exercise and intensive vascular risk reduction in preventing dementia (rrAD Study). Contemp Clin Trials. 2019;79:44–54.

6. Su J, Huang Q, Ren S, Xie F, Zhai Y, Guan Y, et al. Altered Brain Glucose Metabolism Assessed by (18)F-FDG PET Imaging Is Associated with the Cognitive Impairment of CADASIL. Neuroscience. 2019;417:35–44.

7. Rankin KP, Toller G, Gavron L, La Joie R, Wu T, Shany-Ur T, et al. Social Behavior Observer Checklist: Patterns of Spontaneous Behaviors Differentiate Patients With Neurodegenerative Disease From Healthy Older Adults. Front Neurol. 2021;12:683162.

8. Zheng W, Su Z, Liu X, Zhang H, Han Y, Song H, et al. Modulation of functional activity and connectivity by acupuncture in patients with Alzheimer disease as measured by resting-state fMRI. PLoS One. 2018;13(5):e0196933.

9. Verfaillie SC, Tijms B, Versteeg A, Benedictus MR, Bouwman FH, Scheltens P, et al. Thinner temporal and parietal cortex is related to incident clinical progression to dementia in patients with subjective cognitive decline. Alzheimers Dement (Amst). 2016;5:43–52.

10. Tang Y, Xing Y, Zhu Z, He Y, Li F, Yang J, et al. The effects of 7-week cognitive training in patients with vascular cognitive impairment, no dementia (the Cog-VACCINE study): A randomized controlled trial. Alzheimers Dement. 2019;15(5):605–14.

11. Rubinov M, Sporns O. Complex network measures of brain connectivity: uses and interpretations. Neuroimage. 2010;52(3):1059–69.

12. Wind AW, Schellevis FG, Van Staveren G, Scholten RJ, Jonker C, Van Eijk JTM. Limitations of the Mini-Mental State Examination in diagnosing dementia in general practice. International journal of geriatric psychiatry. 1997;12(1):101–8.

13. Ersche KD, Williams GB, Robbins TW, Bullmore ET. Meta-analysis of structural brain abnormalities associated with stimulant drug dependence and neuroimaging of addiction vulnerability and resilience. Curr Opin Neurobiol. 2013;23(4):615–24.

14. Takamiya A, Vande Casteele T, Koole M, De Winter FL, Bouckaert F, Van den Stock J, et al. Lower regional gray matter volume in the absence of higher cortical amyloid burden in late-life depression. Sci Rep. 2021;11(1):15981.

15. Suckling J, Nestor LJ. The neurobiology of addiction: the perspective from magnetic resonance imaging present and future. Addiction. 2017;112(2):360–9.

16. Ferreira LK, Diniz BS, Forlenza OV, Busatto GF, Zanetti MV. Neurostructural predictors of Alzheimer’s disease: a meta-analysis of VBM studies. Neurobiol Aging. 2011;32(10):1733–41.

17. Wu X, Wu Y, Geng Z, Zhou S, Wei L, Ji GJ, et al. Asymmetric Differences in the Gray Matter Volume and Functional Connections of the Amygdala Are Associated With Clinical Manifestations of Alzheimer’s Disease. Front Neurosci. 2020;14:602.

18. Kiesow H, Uddin LQ, Bernhardt BC, Kable J, Bzdok D. Dissecting the midlife crisis: disentangling social, personality and demographic determinants in social brain anatomy. Commun Biol. 2021;4(1):728.

19. Shiino A, Watanabe T, Maeda K, Kotani E, Akiguchi I, Matsuda M. Four subgroups of Alzheimer’s disease based on patterns of atrophy using VBM and a unique pattern for early onset disease. Neuroimage. 2006;33(1):17–26.

20. Zhang Y, Liu Z, Zhao Y. Impulsivity, Social Support and Depression Are Associated With Latent Profiles of Internet Addiction Among Male College Freshmen. Front Psychiatry. 2021;12:642914.

21. Woodworth DC, Sheikh-Bahaei N, Scambray KA, Phelan MJ, Perez-Rosendahl M, Corrada MM, et al. Dementia is associated with medial temporal atrophy even after accounting for neuropathologies. Brain Commun. 2022;4(2):fcac052.

22. Wang WY, Yu JT, Liu Y, Yin RH, Wang HF, Wang J, et al. Voxel-based meta-analysis of grey matter changes in Alzheimer’s disease. Transl Neurodegener. 2015;4:6.

23. He Q, Turel O, Bechara A. Brain anatomy alterations associated with Social Networking Site (SNS) addiction. Sci Rep. 2017;7:45064.

24. Connolly CG, Bell RP, Foxe JJ, Garavan H. Dissociated grey matter changes with prolonged addiction and extended abstinence in cocaine users. PLoS One. 2013;8(3):e59645.

25. Ko CH, Hsieh TJ, Wang PW, Lin WC, Yen CF, Chen CS, et al. Altered gray matter density and disrupted functional connectivity of the amygdala in adults with Internet gaming disorder. Prog Neuropsychopharmacol Biol Psychiatry. 2015;57:185–92.

26. Skouras S, Torner J, Andersson P, Koush Y, Falcon C, Minguillon C, et al. Earliest amyloid and tau deposition modulate the influence of limbic networks during closed-loop hippocampal downregulation. Brain. 2020;143(3):976–92.

27. Younan D, Wang X, Casanova R, Barnard R, Gaussoin SA, Saldana S, et al. PM2.5 associated with gray matter atrophy reflecting increased Alzheimers risk in older women. Neurology. 2020.

28. Alexander-Bloch A, Giedd JN, Bullmore E. Imaging structural co-variance between human brain regions. Nat Rev Neurosci. 2013;14(5):322–36.

29. Cui Y, Liu Y, Yang C, Cui C, Jing D, Zhang X, et al. Brain structural and functional anomalies associated with simultanagnosia in patients with posterior cortical atrophy. Brain Imaging Behav. 2022;16(3):1148–62.

30. Frisoni G, Testa C, Zorzan A, Sabattoli F, Beltramello A, Soininen H, et al. Detection of grey matter loss in mild Alzheimer’s disease with voxel based morphometry. Journal of Neurology, Neurosurgery & Psychiatry. 2002;73(6):657–64.

31. Célestine M, Jacquier-Sarlin M, Borel E, Petit F, Perot J-B, Hérard A-S, et al. Long term worsening of Alzheimer pathology and clinical outcome by a single inoculation of mutated beta-amyloid seeds. 2022.

32. Foo H, Mather KA, Jiang J, Thalamuthu A, Wen W, Sachdev PS. Genetic influence on ageing-related changes in resting-state brain functional networks in healthy adults: A systematic review. Neurosci Biobehav Rev. 2020;113:98–110.

33. Li S, Daamen M, Scheef L, Gaertner FC, Buchert R, Buchmann M, et al. Abnormal Regional and Global Connectivity Measures in Subjective Cognitive Decline Depending on Cerebral Amyloid Status. J Alzheimers Dis. 2021;79(2):493–509.

34. Walker KA, Gross AL, Moghekar AR, Soldan A, Pettigrew C, Hou X, et al. Association of peripheral inflammatory markers with connectivity in large-scale functional brain networks of non-demented older adults. Brain Behav Immun. 2020;87:388–96.

35. Blujus JK, Korthauer LE, Awe E, Frahmand M, Driscoll I. Single Nucleotide Polymorphisms in Alzheimer’s Disease Risk Genes Are Associated with Intrinsic Connectivity in Middle Age. J Alzheimers Dis. 2020;78(1):309–20.

36. Li T, Pappas C, Le ST, Wang Q, Klinedinst BS, Larsen BA, et al. APOE, TOMM40, and sex interactions on neural network connectivity. Neurobiol Aging. 2022;109:158–65.

37. Wang Z, Dai Z, Shu H, Liao X, Yue C, Liu D, et al. APOE Genotype Effects on Intrinsic Brain Network Connectivity in Patients with Amnestic Mild Cognitive Impairment. Sci Rep. 2017;7(1):397.

38. Pietzuch M, Bindoff A, Jamadar S, Vickers JC. Interactive effects of the APOE and BDNF polymorphisms on functional brain connectivity: the Tasmanian Healthy Brain Project. Sci Rep. 2021;11(1):14514.

39. Contreras JA, Aslanyan V, Sweeney MD, Sanders LMJ, Sagare AP, Zlokovic BV, et al. Functional connectivity among brain regions affected in Alzheimer’s disease is associated with CSF TNF-alpha in APOE4 carriers. Neurobiol Aging. 2020;86:112–22.

40. Soman SM, Raghavan S, Rajesh PG, Mohanan N, Thomas B, Kesavadas C, et al. Does resting state functional connectivity differ between mild cognitive impairment and early Alzheimer’s dementia? J Neurol Sci. 2020;418:117093.

41. Zhou B, Liu Y, Zhang Z, An N, Yao H, Wang P, et al. Impaired functional connectivity of the thalamus in Alzheimer’s disease and mild cognitive impairment: a resting-state fMRI study. Current Alzheimer Research. 2013;10(7):754–66.

42. Zhao Q, Lu H, Metmer H, Li WX, Lu J. Evaluating functional connectivity of executive control network and frontoparietal network in Alzheimer’s disease. Brain research. 2018;1678:262–72.

43. Mohammadian F, Zare Sadeghi A, Noroozian M, Malekian V, Abbasi Sisara M, Hashemi H, et al. Quantitative Assessment of Resting-State Functional Connectivity MRI to Differentiate Amnestic Mild Cognitive Impairment, Late-Onset Alzheimer’s Disease From Normal Subjects. Journal of Magnetic Resonance Imaging. 2023;57(6):1702–12.

44. Shi JY, Wang P, Wang BH, Xu Y, Chen X, Li HJ. Brain Homotopic Connectivity in Mild Cognitive Impairment APOE-epsilon4 Carriers. Neuroscience. 2020;436:74–81.

45. Yamazaki Y, Zhao N, Caulfield TR, Liu CC, Bu G. Apolipoprotein E and Alzheimer disease: pathobiology and targeting strategies. Nat Rev Neurol. 2019;15(9):501–18.

46. Lin H, Li M, Zhan Y, Lin L, Yang K, Hu S, et al. Disrupted white matter functional connectivity in aMCI APOEepsilon4 carriers: a resting-state study. Brain Imaging Behav. 2021;15(4):1739–47.

47. Kulkarni P, Grant S, Morrison TR, Cai X, Iriah S, Kristal BS, et al. Characterizing the human APOE epsilon 4 knock-in transgene in female and male rats with multimodal magnetic resonance imaging. Brain Res. 2020;1747:147030.

48. Tahmi M, Rippon B, Palta P, Soto L, Ceballos F, Pardo M, et al. Brain Amyloid Burden and Resting-State Functional Connectivity in Late Middle-Aged Hispanics. Front Neurol. 2020;11:529930.

49. Cruchaga C, Chakraverty S, Mayo K, Vallania FL, Mitra RD, Faber K, et al. Rare variants in APP, PSEN1 and PSEN2 increase risk for AD in late-onset Alzheimer’s disease families. PloS one. 2012;7(2):e31039.

50. Chan E, Altendorff S, Healy C, Werring DJ, Cipolotti L. The test accuracy of the Montreal Cognitive Assessment (MoCA) by stroke lateralisation. Journal of the neurological sciences. 2017;373:100–4.

51. Smith T, Gildeh N, Holmes C. The Montreal Cognitive Assessment: validity and utility in a memory clinic setting. The Canadian Journal of Psychiatry. 2007;52(5):329–32.

52. Cheah WK, Teh HL, Huang DXH, Ch’ng ASH, Choy MP, Teh EE, et al. Validation of Malay version of Montreal cognitive assessment in patients with cognitive impairment. Clin Med Res. 2014;3(3):56–60.

53. Mf F. Folstein SE. McHugh PR.“Mini-mental state.” A practical method for grading the cognitive state of patients for the clinician. J Psychiatr Res. 1975;12(3):189–98.

54. Ibrahim NM, Shohaimi S, Chong H-T, Rahman AHA, Razali R, Esther E, et al. Validation study of the Mini-Mental State Examination in a Malay-speaking elderly population in Malaysia. Dementia and geriatric cognitive disorders. 2009;27(3):247–53.

55. Stenger JE, Lobachev KS, Gordenin D, Darden TA, Jurka J, Resnick MA. Biased distribution of inverted and direct Alus in the human genome: implications for insertion, exclusion, and genome stability. Genome research. 2001;11(1):12–27.

56. Teh HL, Suan M, Azri M, Ahmad R, Yahya MH. Development and validation of Dementia Solat Score for detecting cognitive impairment among Muslim patients: A pilot study. Neurology Asia. 2021;26(4).

57. Ashburner J, Friston KJ. Voxel-based morphometry--the methods. Neuroimage. 2000;11(6 Pt 1):805–21.

58. Pando-Naude V, Toxto S, Fernandez-Lozano S, Parsons CE, Alcauter S, Garza-Villarreal EA. Gray and white matter morphology in substance use disorders: a neuroimaging systematic review and meta-analysis. Transl Psychiatry. 2021;11(1):29.

59. Hobson J. The montreal cognitive assessment (MoCA). Occupational Medicine. 2015;65(9):764–5.

60. Morris JC, Edland S, Clark C, Galasko D, Koss E, Mohs R, et al. The Consortium to Establish a Registry for Alzheimer’s Disease (CERAD): Part IV. Rates of cognitive change in the longitudinal assessment of probable Alzheimer’s disease. Neurology. 1993;43(12):2457-.

61. Torralva T, Gleichgerrcht E, Ibañez A, Manes F. The frontal lobes. Oxford Textbook of Cognitive Neurology and Dementia Oxford University Press, UK. 2016:27–38.

62. Pastore A. Individual decision making, reinforcement learning and myopic behaviour: University of Sheffield; 2019.

63. Abedelahi A, Hasanzadeh H, Hadizadeh H, Joghataie MT. Morphometric and volumetric study of caudate and putamen nuclei in normal individuals by MRI: effect of normal aging, gender and hemispheric differences. Polish journal of radiology. 2013;78(3):7.

64. Japee S, Holiday K, Satyshur MD, Mukai I, Ungerleider LG. A role of right middle frontal gyrus in reorienting of attention: a case study. Frontiers in systems neuroscience. 2015;9:23.

65. Li HJ, Hou XH, Liu HH, Yue CL, He Y, Zuo XN. Toward systems neuroscience in mild cognitive impairment and Alzheimer’s disease: A meta-analysis of 75 fMRI studies. Human brain mapping. 2015;36(3):1217–32.

66. Colangeli S, Boccia M, Verde P, Guariglia P, Bianchini F, Piccardi L. Cognitive reserve in healthy aging and Alzheimer’s disease: a meta-analysis of fMRI studies. American Journal of Alzheimer’s Disease & Other Dementias®. 2016;31(5):443–9.

67. Fortel I, Korthauer LE, Morrissey Z, Zhan L, Ajilore O, Wolfson O, et al. Connectome Signatures of Hyperexcitation in Cognitively Intact Middle-Aged Female APOE-epsilon4 Carriers. Cereb Cortex. 2020;30(12):6350–62.

68. Mevel K, Chételat G, Eustache F, Desgranges B. The default mode network in healthy aging and Alzheimer’s disease. International journal of Alzheimer’s disease. 2011;2011.

