## Supplementary figures and images for "Apolipoprotein E-Genotyping and MRI Study for Alzheimer’s Disease Classification: PCR-RFLP and Restricted Enzymes AfIII for RS429358 and HaeII for RS7412"

### Table 1

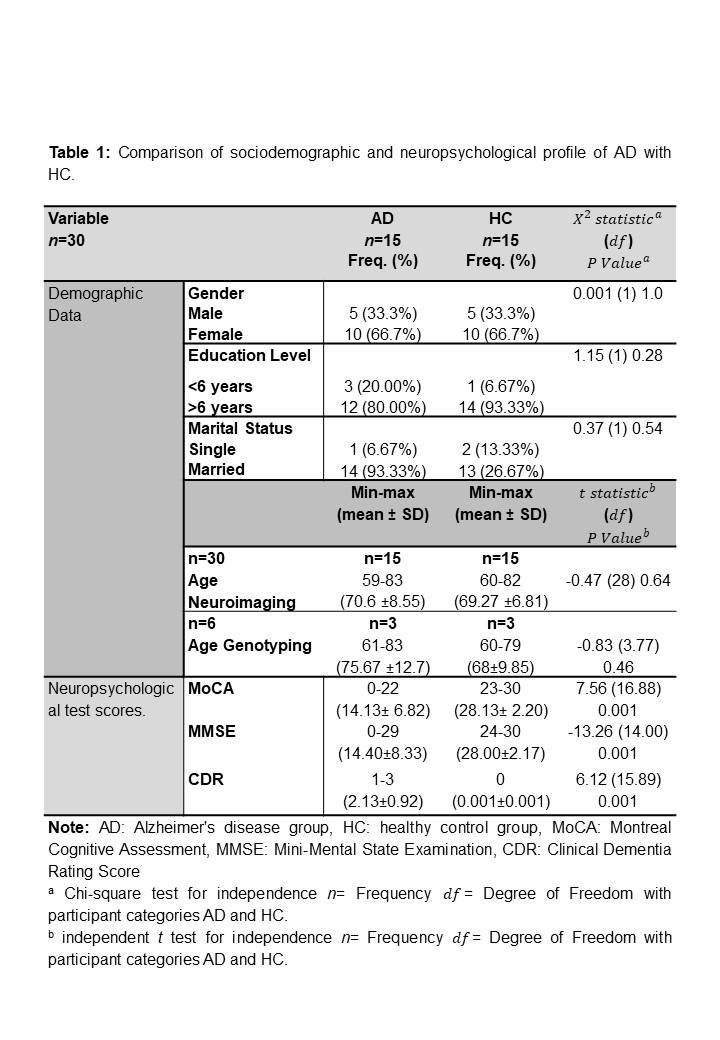

### Table 2

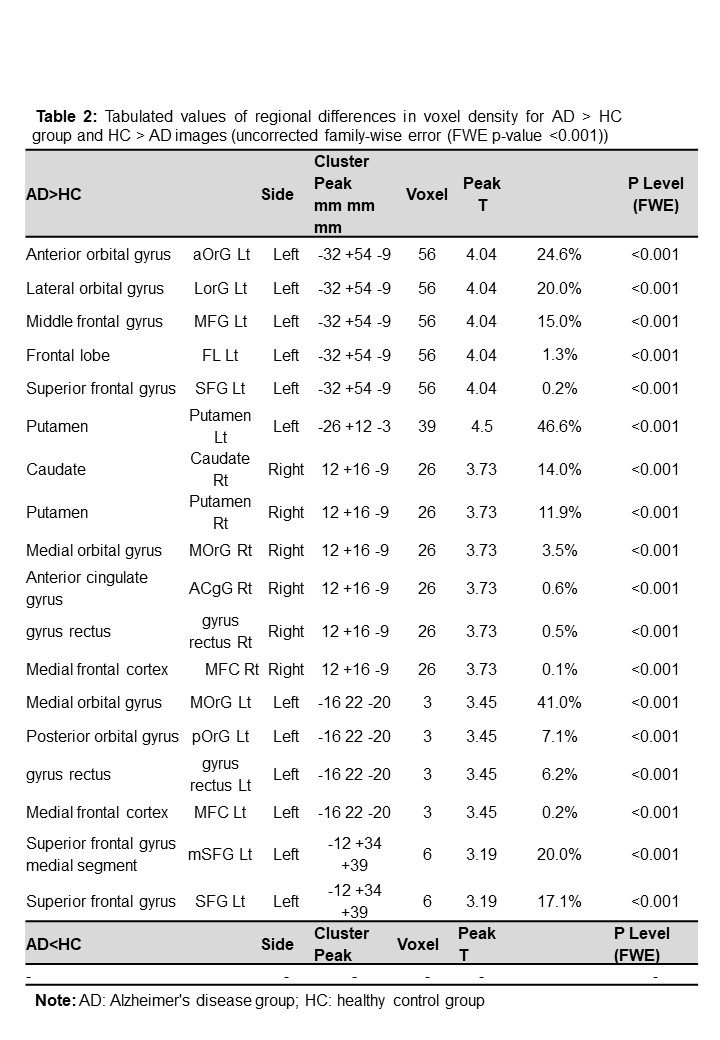

### Table 3

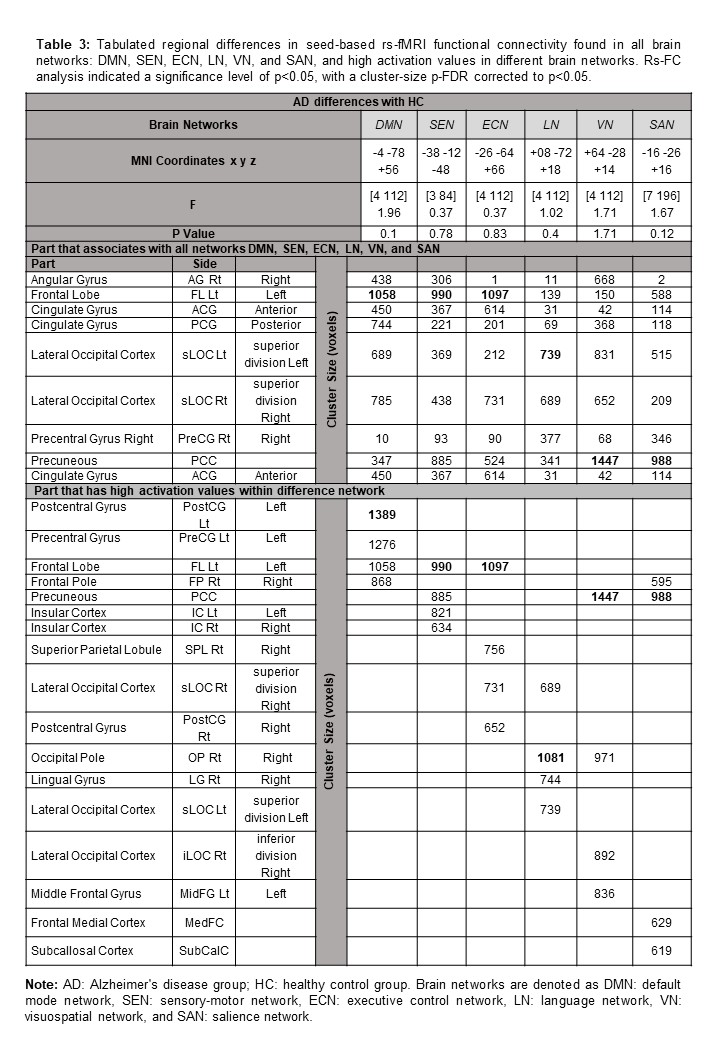
